# Multi-omics data integration identifies novel biomarkers and patient subgroups in inflammatory bowel disease

**DOI:** 10.1101/2024.07.23.24310846

**Authors:** António José Preto, Shaurya Chanana, Daniel Ence, Matt D. Healy, Daniel Domingo-Fernández, Kiana A. West

**Affiliations:** Enveda Biosciences, 5700 Flatiron Parkway, Boulder, CO, 80301, USA

## Abstract

**Objective:** In this work, we explored one of the largest multi-omics cohorts in Inflammatory Bowel Disease (IBD), the Study of a Prospective Adult Research Cohort (SPARC IBD), with the goal of identifying predictive biomarkers for Crohn’s Disease (CD) and Ulcerative Colitis (UC) and elucidating patient subtypes.

**Design:** We analyzed genomics, transcriptomics (gut biopsy samples), and proteomics (blood plasma) from hundreds of patients from SPARC IBD. We trained a machine learning model that classifies UC vs. CD samples. In parallel, we leveraged multi-omics data integration to unveil patient subgroups in each of the two indications independently and analyzed the molecular phenotypes of these patient subpopulations.

**Results:** The high performance of the model showed that multi-omics signatures are able to discriminate between the two indications. The most predictive features of the model, both known and novel omics signatures for IBD, can potentially be used as diagnostic biomarkers. Patient subgroups analysis in each indication uncovered omics features associated with disease severity in UC patients, and with tissue inflammation in CD patients. This culminates with the observation of two CD subpopulations characterized by distinct inflammation profiles.

**Conclusion:** Our work unveiled potential biomarkers to discriminate between CD and UC and to stratify each population into well-defined subgroups, offering promising avenues for the application of precision medicine strategies.

## 1. Introduction

Inflammatory Bowel Disease (IBD) encompasses Crohn’s Disease (CD) and Ulcerative Colitis (UC), both of which are chronic and complex conditions that impact the gastrointestinal tract. Characterized by its heterogeneity, IBD presents a spectrum of disease manifestations, including variability in disease location within the gastrointestinal tract, disease behaviors, and levels of disease activity among patients (Baumgart and Carding, 2007). Despite significant research efforts aimed at unraveling the disease’s complexity—ranging from identifying genetic variants and environmental factors to analyzing the gut microbiome—the pathophysiological mechanisms defining the distinct clinical subtypes of IBD remain partially understood. The challenge lies in pinpointing the specific molecular pathways that contribute to this heterogeneity and developing targeted treatment approaches.

The emergence of multi-omics technologies has opened new pathways for understanding the intricate biological networks at play in IBD (Weersma *et al*., 2018; Lloyd-Price *et al*., 2019; Agrawal *et al*., 2022). By generating and analyzing genomics, transcriptomics, proteomics, and metabolomics data, researchers now can better understand the molecular characteristics of IBD (Subramanian et al., 2020). Several projects have been initiated to amass patient-level omics data, demonstrating the commitment to this comprehensive approach (Sudhakar *et al.,* 2022). Notable examples include the 1000IBD project, which has collected genomics, gut microbiome taxonomic profiles, and fecal metagenomics from over 1,000 patients (Imhann *et al*., 2019), and the IBD Database, featuring a wide range of omics data from more than 100 patients (Lloyd-Price *et al*., 2019). Additionally, the International Inflammatory Bowel Disease Genetics Consortium (IIBDGC) focuses on genetic data from thousands of patients globally. Yet many of these initiatives have primarily concentrated on metagenomics and genomics, often overlooking proteomics and transcriptomics from blood and biopsies. The recent SPARC IBD cohort aims to bridge this gap by enrolling over 3,000 CD and UC patients and incorporating a diverse array of omics data, thus paving the way for groundbreaking insights into the disease (Raffals *et al.,* 2022).

Leveraging such cohorts, research efforts have focused on identifying biomarker signatures for IBD. For instance, Mo and colleagues (2023) used transcriptomics data from 357 UC patients to cluster them into three subpopulations that correlated with clinical responses. Similarly, Janker and colleagues (2023) examined proteomics and metabolomics in blood plasma and colon tissue from UC patients, those in remission, and controls, revealing upregulation of inflammatory pathways in active UC, which normalize upon remission. This analysis utilized mixOmics and Gaussian modeling, though the study’s impact was limited by its small sample size. In a broader approach, Motwani *et al*. (2024) utilized the SPARC dataset to examine the correlation between non-invasive biomarkers (CRP and FCP) and patient-reported outcomes (PROs), discovering that while there is a correlation, it varies by disease location and type.

In this work, we leveraged the largest multi-omics IBD dataset available in pursuit of novel, clinically-relevant biomarkers for diagnostics and patient stratification. We used machine learning (ML) to empirically determine multiple omics features that distinguish UC from CD patients, resulting in potential biomarkers that may aid diagnosis of patients with indeterminate colitis. In parallel, we integrated multiple omics layers to identify and characterize patient subpopulations based on clinical phenotypes such as disease severity and intestinal inflammation.

## 2. Methods

The results published here are based on data obtained from the IBD Plexus program of the Crohn’s & Colitis Foundation. **Figure 1** outlines the methodology of this work and the structure of the methods section. In the first subsection, we describe the SPARC IBD cohort and the three omics modalities used (**Figure 1 left)**. The following two sections outline the multi-omics data preparation and processing (**Figure 1 center)**. Finally, in the last two subsections, we describe the supervised machine learning classifier and the unsupervised method known as Multi-Omics Factor Analysis (MOFA) used in this work (**Figure 1 right)**.

**Figure 1.**
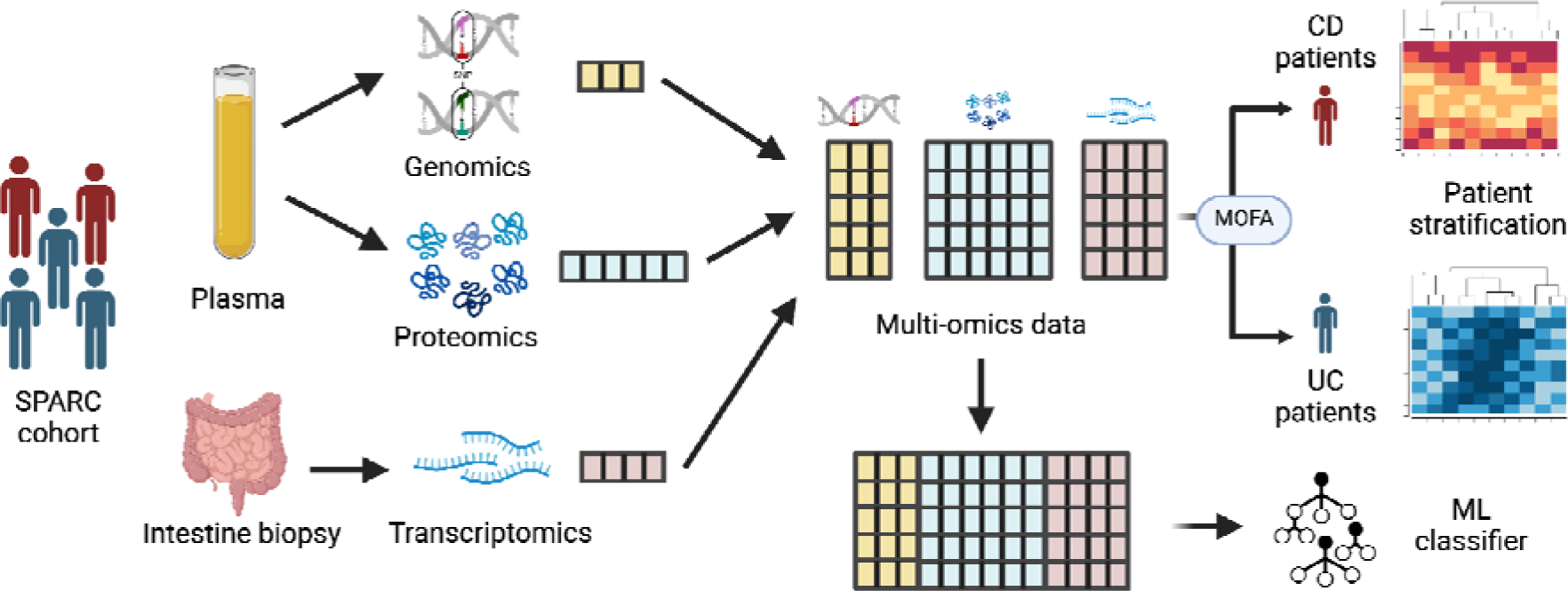
Outline of the methodology. The starting point of our work are three types of omics data (proteomics, genomics, and transcriptomics) generated from samples from IBD patients (i.e., CD and UC). We processed these three omics modalities to generate a multi-omics dataset. In one venue, we combined the omics data and trained a ML classifier that can accurately differentiate between samples derived from UC and CD patients. Parallely, we explored the characteristics of different subpopulations in each indication (CD and UC) using Multi Omics Factor Analysis (MOFA). This figure was created by usin BioRender.com.

### 2.1. SPARC IBD cohort

The starting point for this research is the Study of a Prospective Adult Research Cohort (SPARC IBD), a component of the Crohn’s & Colitis Foundation’s IBD Plexus research platform. The SPARC IBD cohort as of April 2024, contained multiple data modalities across different timepoints for several thousands of patients with IBD. For the purpose of this work, we subset the full cohort to samples of patients characterized with three omics modalities: genomics, transcriptomics, and proteomics **(Supplementary Figure 1)**. **Table 1** summarizes the different batches across these three omics modalities for patients with all three omics modalities available. We refer to **Supplementary Text 1** and (Raffals *et al.,* 2022) for details on the experimental data preparation. Additionally, **Supplementary Table 2** describes the patient demographics.

**Table 1.** Sample batches in each of the different omics modalities for patients with all three omics modalities available.

| SPARC IBD Cohort Batch | Modality | Unique patients |
| --- | --- | --- |
| sparc-genotyping | Genomics | 537 |
| sparc-genewiz-2021 | Transcriptomics | 165 |
| sparc-cd-genewiz | Transcriptomics | 111 |
| sparc-genewiz | Transcriptomics | 117 |
| sparc-genewiz-2022 | Transcriptomics | 207 |
| sparc-olink | Proteomics | 537 |

### 2.2. Data processing

Transcripts without a gene name were dropped. Using PyDESeq2 (Muzellec *et al.,* 2023) we normalized each of the batches and multiplied by the size factors (i.e., related to the number of reads in the library), thus making the expression values independent from the number of reads. We dropped features with zero variance (i.e., same value across all samples).

We deployed multiple standard approaches to minimize the batch effect observed in the transcriptomics data, such as pyComBat (Behdenna *et al.,* 2023) and linear modeling with limma (Ritchie *et al.,* 2015). However, this step alone was unable to successfully minimize the batch effect; batch remained the main source of variance for numerous transcriptomics features. To mitigate this technical variation, we added an additional step to the batch effect correction approach. For each of the features, we calculated the average of the variance for each of the batches (henceforth, intra-batch variance) and dropped the features where the intra-batch variance was lower than 0.05. We performed batch correction on the remaining features using pyComBat. We processed genomics data using VCFtools (Danecek *et al.,* 2011) by reducing the raw data to biallelic sites with a minor allele frequency of at least 0.05. We annotated the data using PyEnsembl, with the Ensembl 110 release of the human genome (GRCh38.p14) of the human database (Martin *et al.,* 2023). Proteomics data were used as provided without features tagged as low quality (i.e., “QC warning”) and QC samples.

Lastly, we matched all three omics for each patient by collection date with a threshold of 7 days **(details in Supplementary Text 2)**. After collapsing the metadata, we arrive at a total of 1,431 patients with transcriptomics data, 1,563 patients with proteomics data, 1,641 patients with genomics array information and 1,641 patients with genomics exome information. Hereafter, we will refer to the genomics information solely as the genomics array information. In the intersection of the three omics types, we have 857 unique patients **(Supplementary** Figure 2). For the following analyses, we exclusively considered samples from these patients with all three omics.

Regarding the features, for each omics type, and after removing the features with null variance, we arrived at 166,916 for genomics, 19,442 for transcriptomics and 2,940 for proteomics.

### 2.3. Building an ML model to classify UC vs. CD

We trained an XGBoost classifier (Chen and Guestrin, 2016) to predict whether the omics data originated from UC or CD patients (CD=703 and UC=320) **(Supplementary Figure 3).** We chose this model as it is considered to be state-of-the-art in generic tabular data as well as on similar large biomedical cohorts (Nielsen *et al.,* 2024). Furthermore, while neural network architectures could have been used, we preferred XGBoost due its inherent interpretability, and its aforementioned consistent high performance in similar data.

To evaluate the performance of the model, we first performed an 80/20 split on the unique patient identifiers, generating independent train and test sets. We normalized the train and test data by subtracting the mean and scaling by the variance of the training data. We trained the model using a nested 5-fold stratified cross-validation on the training set. Specifically, we trained a hyperparameter-optimized model on each of five train-validation splits. Parameters for each model were optimized using five-fold cross-validation exclusively within the training set of each fold to avoid overfitting. Splits were stratified to equalize the relative proportions of UC and CD patients. The hyperparameters used and their ranges are shown in **Supplementary Table 1.** We evaluate the 5 models on the test and validation sets, in order to deliver the performance results by metric and its respective standard deviation across the models. The performance evaluation metrics used in this categorical prediction setting are: accuracy, F1-score, AUR-ROC, precision, and recall. Finally, we look at the most predictive features of the model based on gain, which measures the increase in performance brought by a feature in the branches it acts upon.

### 2.4. Multi-omics driven clustering

In order to perform unsupervised patient stratification, we leveraged Multi-Omics Factor Analysis (MOFA) (Argelaguet *et al*. 2018). Our goal when using this technique was simultaneously performing integration of multiple omics data sources and types as well as identifying sets of descriptors that explain biological and physiological functions. Ultimately, we aimed at identifying phenotypic-driven clusters of patient samples that are likely to be more responsive to different therapies.

CD and UC are heterogeneous conditions with multiple subtypes (Selin *et al*., 2021). To account for this, we separately ran MOFA analyses for each diagnosis (CD and UC). Considering the specificity of transcriptomics patterns in different tissues (Figueiredo *et al*., 2022), which we observe later on in our analyses **(subsection 3.1)**, we focused solely on the colon samples for UC and modeled colon samples separately from small intestine samples for CD. We independently standardized the omic data features by removing the mean and scaling to unit variance prior to modeling. We refer to **Supplementary Text 3** for more details about MOFA and the parameters used.

We identify the factors of interest by their relationship to either macroscopic appearance (intestinal inflammation based on the biopsy sampled) or disease severity. The factors are a result of the weighted contributions of features that are likely to be involved in similar biological functions (Argelaguet *et al*. 2018). From these factors, and for each omics type, we retrieve the omics signatures with the highest absolute weights for that factor. On this selection of features, we conducted pathway analysis using GSEApy (Fang *et al*. 2023) with the gene sets from GO Biological Process (2023), considering enriched pathways as the ones with an adjusted *p*-value less than 0.01. Pathways were ranked according to the combined score returned by EnrichR (Chen *et al.,* 2013).

## 3. Results

### 3.1. Correcting for batch effect unlocks the power of multi-omics data

We found that the transcriptomics data exhibited a strong batch effect that was the predominant source of variation and hindered identifying any biological patterns on the data **(Figure 2A)**. After applying ComBat, the batch effect had drastically diminished, from a silhouette score of 0.28 on the PCA plot **(Figure 2A)** to −0.05, after correction (**Figure 2D**).

**Figure 2.**
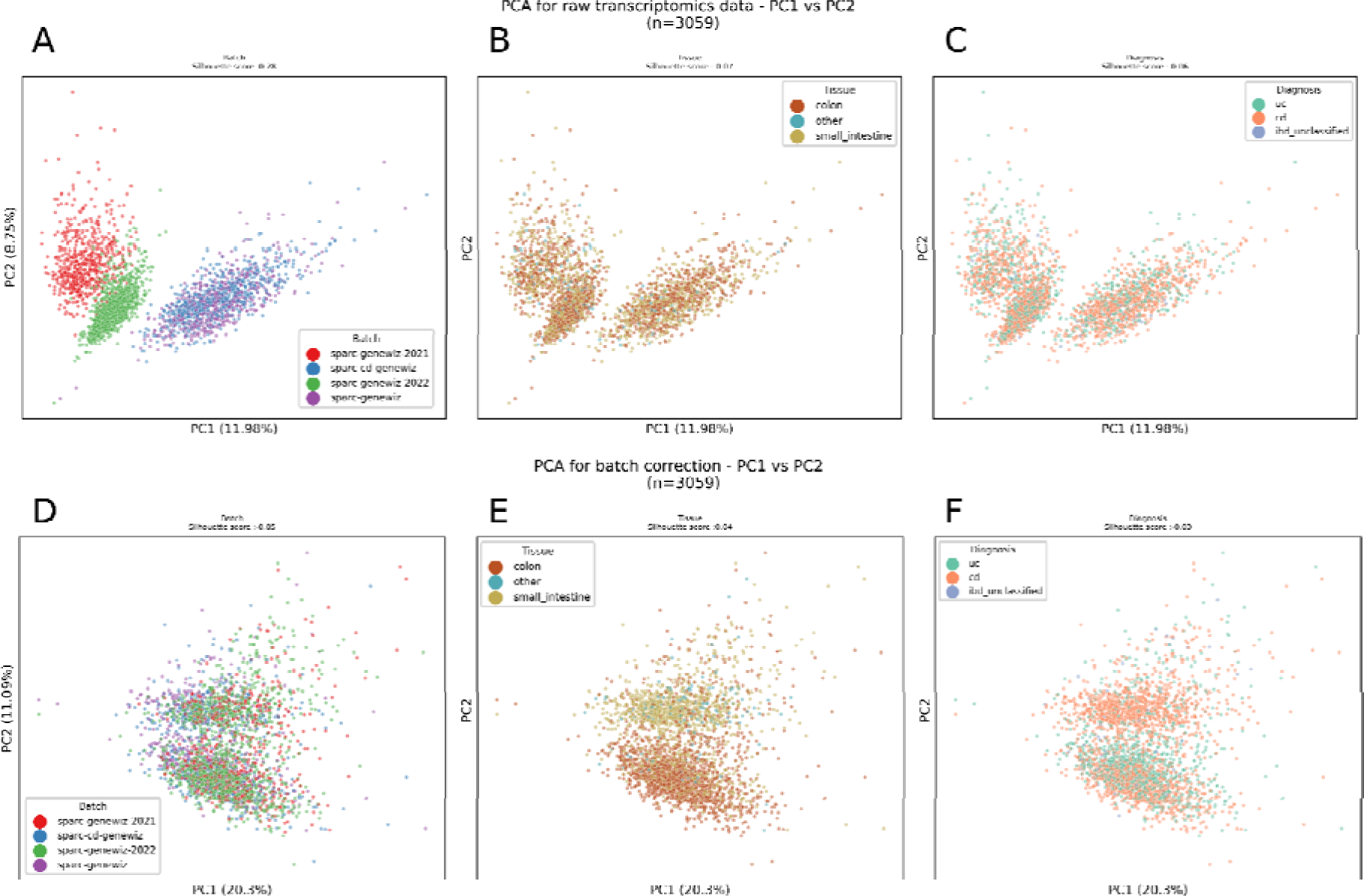
First row: PCA of the first two components of the transcriptomics samples, colored by batch **(A)**, tissue **(B)** and diagnosis **(C)**. Second row: PCA of the first two components of the transcriptomics samples after correcting for batch effect using batch variance and pyComBat, colored by batch **(D)**, tissue **(E)** and diagnosis **(F)**.

This is further corroborated by calculating the Mutual Information (MI) scores (Yu *et al*. 2023) between the features and the batch. Prior to batch correction, 22.28% of the transcriptomics features had MI scores above 0.2; afterwards, this percentage dropped to 3.55% (**Supplementary Figure 4**). After correcting the batch effect, the PCA revealed a slight association between the samples and tissue (**Figure 2E**), which is expected given that transcription patterns are known to be tissue-specific (Figueiredo *et al*., 2022), and most of the samples in the dataset are derived from colon and small intestine. Lastly, other variables such as diagnosis do not show an apparent association in the PCA (**Figure 2C and F**).

### 3.2. Multi-omics signatures can accurately differentiate between CD and UC patients

We explored the differences between UC and CD by training an ML model to classify between both diseases using multi-omics data from each sample as an input. The ML classifier was able to distinguish between the two indications (Accuracy: 0.77±0.02, Precision (CD): 0.77±0.02; Recall (CD): 0.87 ± 0.02; Precision (UC): 0.75±0.02; Recall (UC): 0.57±0.02; AUC: 0.75 ± 0.02), **(Figure 3A and B)**. These results demonstrate that a multi-omics classifier can accurately distinguish between the two phenotypes.

**Figure 3.**
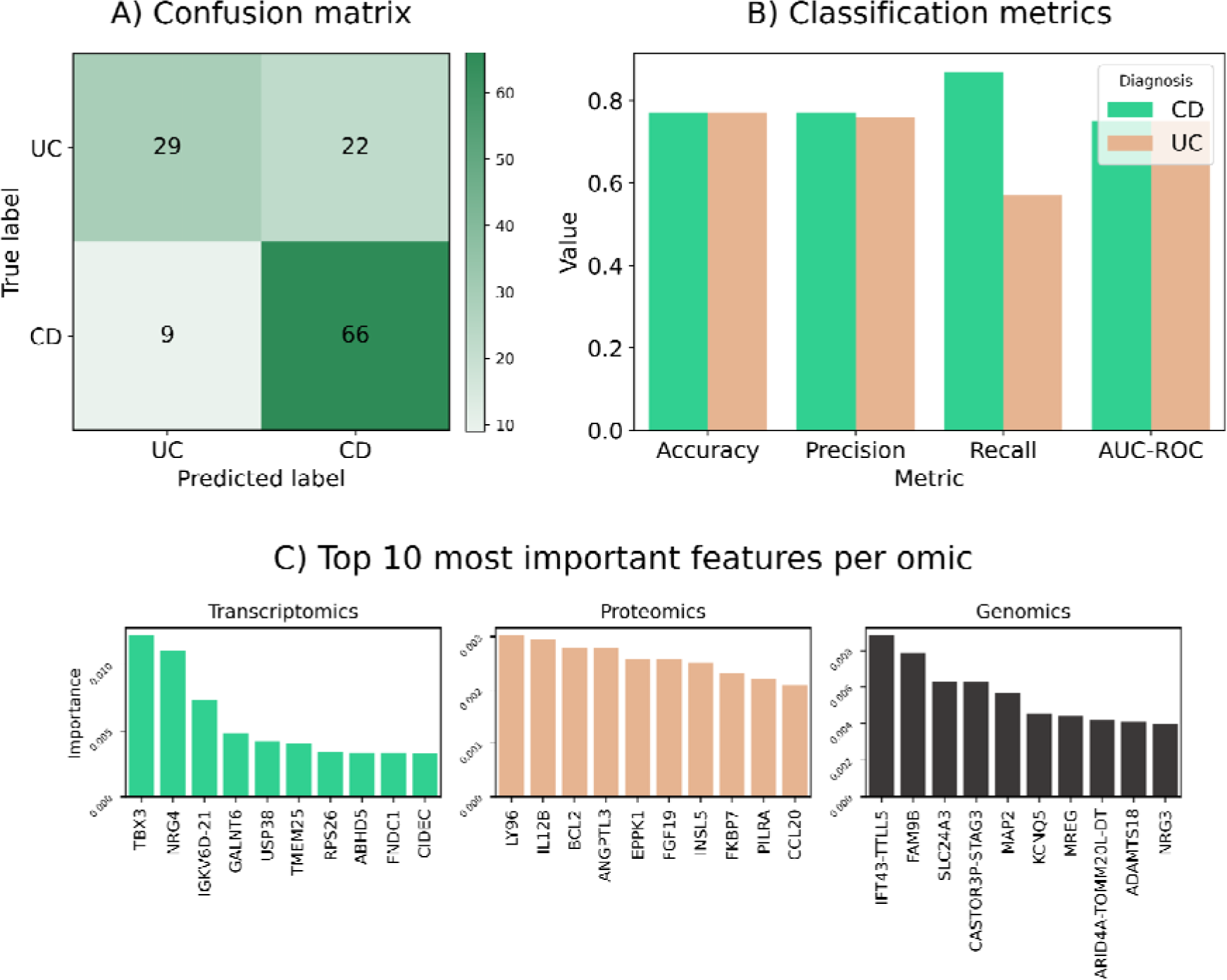
**A)** Confusion matrix of the predictions on the test set of one of the five CVs of the model. **B)** Bar plots with different performance metrics and the standard deviation of the classifier over the five CVs evaluated on the test set. Performance is close to 0.8 across all metrics evaluated. **C)** Top 10 most predictive features in the three omics.

When looking into the most predictive features, we observed a few that stood out from the rest **(Figure 3C)**. Particularly for transcriptomics, there are several known genes associated with inflammation such as Immunoglobulin Kappa Variable 6D-21 (IGKV6D-21) (Hu *et al*. 2023), GALNT6 (Ding *et al*. 2023) TBX3 (Khan *et al*. 2021), NRG4 (Shi *et al*. 2022) and USP38 (Gong *et al*. 2023). The function of these genes varies– IGKV6D-21 has been connected with ischemic stroke and IBD, GALNT6 has been shown to intervene in the NF-κB/NLRP3/GSDMD and GSDME pathways, both related to IBD, NRG4 was linked to endothelial inflammation, and USP38 has been linked to inflammation in the heart. However, our results also revealed that a few of them have not yet been clearly linked with IBD or any of its two phenotypes (UC and CD) such as RPS26 and TMEM25, suggesting they could be novel genes involved in the pathogenesis of these phenotypes. Similarly, FNDC1 has previously been linked to IBD with no clear confirmation (Wuensch *et al*. 2019).

In the case of proteomics, which is measured in plasma, the top proteins ranked by importance are involved in cardiometabolic or inflammatory processes and most of them are reported to be linked with IBD. For example, INSL5, together with other anti-inflammatory cytokines, was reported to differentiate between UC and CD (Skok *et al*. 2021). Similarly, low levels of EGFR are associated with the risk of developing IBD (Yang *et al*., 2024). Furthermore, we find four other inflammation-related proteins: IL12B (Lee *et al*., 2016), FGF19 (Łukawska *et al*., 2024), LY96 (Bank *et al*. 2014), CCL20 (Kaser *et al*. 2004) and BCL2 (Weder *et al*. 2018), which are associated with disease activity. On the other hand, there are other proteins not yet linked with IBD among the top such as ANGPTL3. We do not find loci associated with IBD features within the top genomics features.

### 3.3. Identifying molecular profiles that correlate with disease severity in Ulcerative Colitis

In this subsection, we modeled multi-omics UC samples using MOFA. Our goal was to leverage this unsupervised technique to pinpoint characteristic molecular profiles that correlate with clinical phenotypes such as disease severity (based on endoscopy) and macroscopic appearance (i.e., whether the sample is inflamed or not). By exploring these multi-omics profiles, we ultimately intend to identify biomarkers that can be used to define patient subpopulations.

By clustering the factors from MOFA, we observed a correlation (R^2) of 0.40 between factor 1 and disease severity (**Figure 4A** and **Supplementary Figure 5**). The Kendall’s Tau test further corroborate this, showing a moderate yet significant (*p*-value = 8*10LL) correlation of 0.30. This correlation suggests that the features associated with this factor can potentially be used to distinguish between patients with severe or moderate disease from patients with mild disease or in remission. Thus, we investigated the top 150 features across the three omics based on their weights for this factor, since these top features are responsible for the variability observed within this factor. To identify the features that characterize patients with severe disease, we applied ANOVA to proteomics and transcriptomics features and chi-square to genomics (p-value < 0.01), using severe versus the rest as groups.

**Figure 4.**
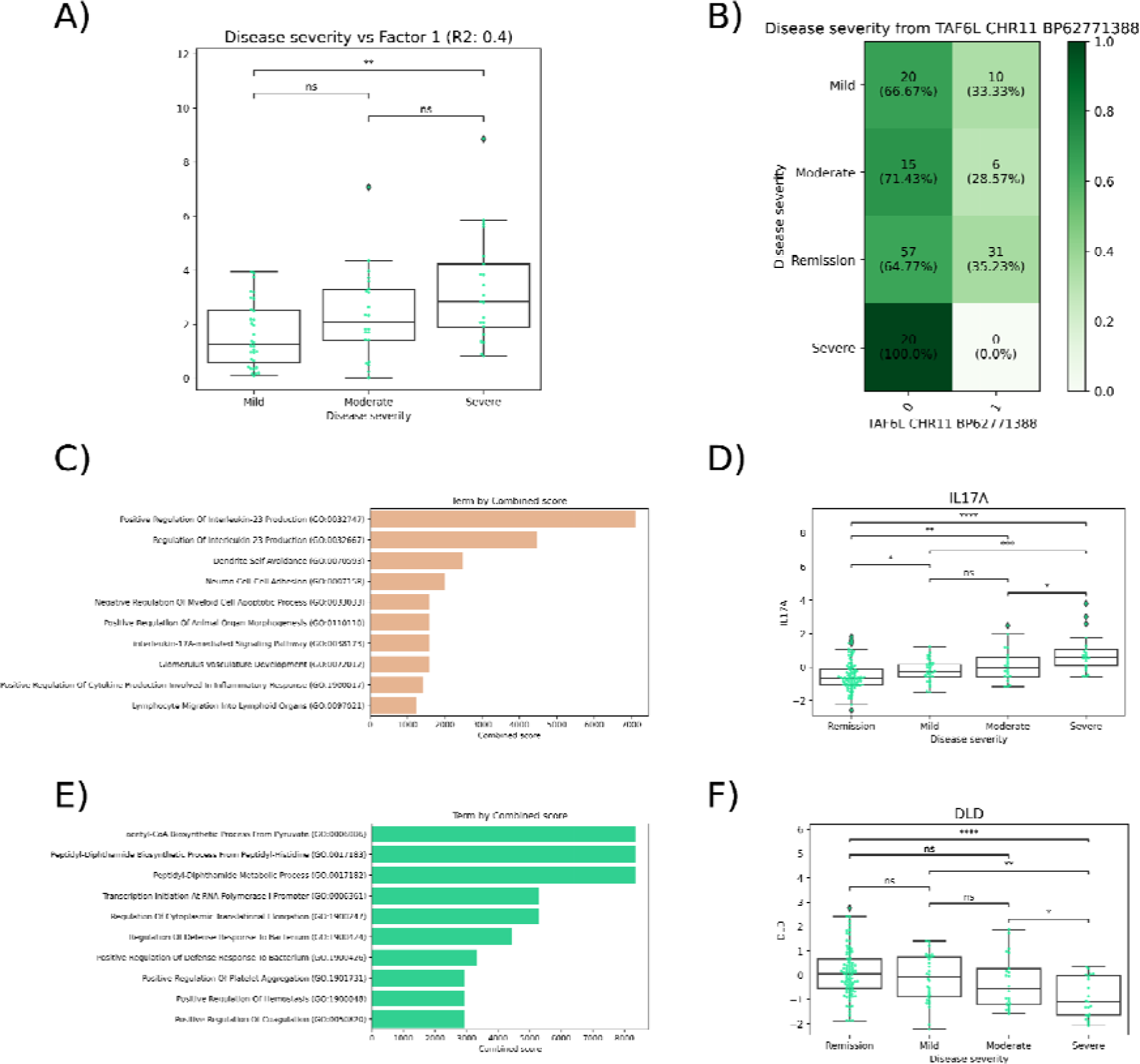
**A)** Distribution of the absolute factor 1 values stratified by disease severity based on endoscopy. The correlation between factor 1 and disease severity is 0.4 considering mild=1, moderate=2, severe=3 to make disease severity a numeric variable. **B)** Distribution of the genotypes (counts and percentage for each row) for TAF6L (chr11_bp62771388) stratified by disease severity based on endoscopy. 0 corresponds to the reference allele and 1 the first alternate allele. **C)** Top 10 enriched pathways using the significant proteomics features (orange). **D)** Distribution of the expression values for IL17A (proteomics). **E)** Top 10 enriched pathways using the significant transcriptomics features (green). **F)** Distribution of the expression values for DLD (transcriptomics).

Among the significant features, we identified several potential biomarkers for disease severity across the three omics. Firstly, for genomics, we found that all severe patients have the reference allele (0) of TAF6L (chr11_bp62771388) **(Figure 4B)**, while ZNF268 (chr12_bp133202004) and CEP164 (chr11_bp117403235) severe patients typically have the first alternate allele (1) (**Supplementary Figure 6)**. In the case of proteomics, we observed that disease severity correlates with IL17A (**Figure 4D)**, EPO, and REG1B (**Supplementary Figure 6)**. Similarly, in transcriptomics, we found several examples such as EMILIN2 and DL, which exhibit a positive and an inverse correlation with disease severity, respectively **(Figure 4F and Supplementary Figure 6)**. Lastly, we investigated the enriched pathways of these significant features by running pathway enrichment analysis on each omics modality independently. Among the enriched pathways, we found interleukin and cytokine signaling pathways for proteomics (**Figure 4C)** and metabolic pathways as well as pathways related to hemostasis and response to pathogens.

### 3.4. Investigating the molecular phenotype of inflamed samples in Crohn’s disease reveals two subpopulations

Similar to the previous section on UC, we modeled multi-omics CD samples using MOFA aimed at identifying molecular profiles that correlate with clinical phenotypes. As mentioned in Methods, we separated colon and small intestine transcriptomics samples, due to the strong tissue effect. Here, we exclusively report the findings derived from colon transcriptomics, since we did not find a clear correlation when the multi-omics samples comprised small intestine transcriptomics instead.

First, we began by performing hierarchical clustering across all samples based on all factors yielded by MOFA to assess whether any factor was correlating with any variable of clinical interest **(Figure 5A)**. While the clusters did not reveal any clear correlation with any of the variables of that could introduce undesired bias such as batch, sex, and age, we found that factor 3, which is the only factor with relevant contribution for more than one omics modality **(Supplementary Figure 7)**, exhibited a clear correlation with macroscopic appearance **(Figure 5B)**. For this factor, non inflamed samples exhibit values of factor 3 close to zero, while the majority of the inflamed samples have either highly positive or highly negative factor 3 values, suggesting two distinct inflammation phenotypes. Notably, inflamed samples tend to have a higher Simple Endoscopic Score for Crohn’s Disease (SES-CD), suggesting an increased disease severity **(Supplementary Figure 8)** (Daperno *et al*., 2004).

**Figure 5.**
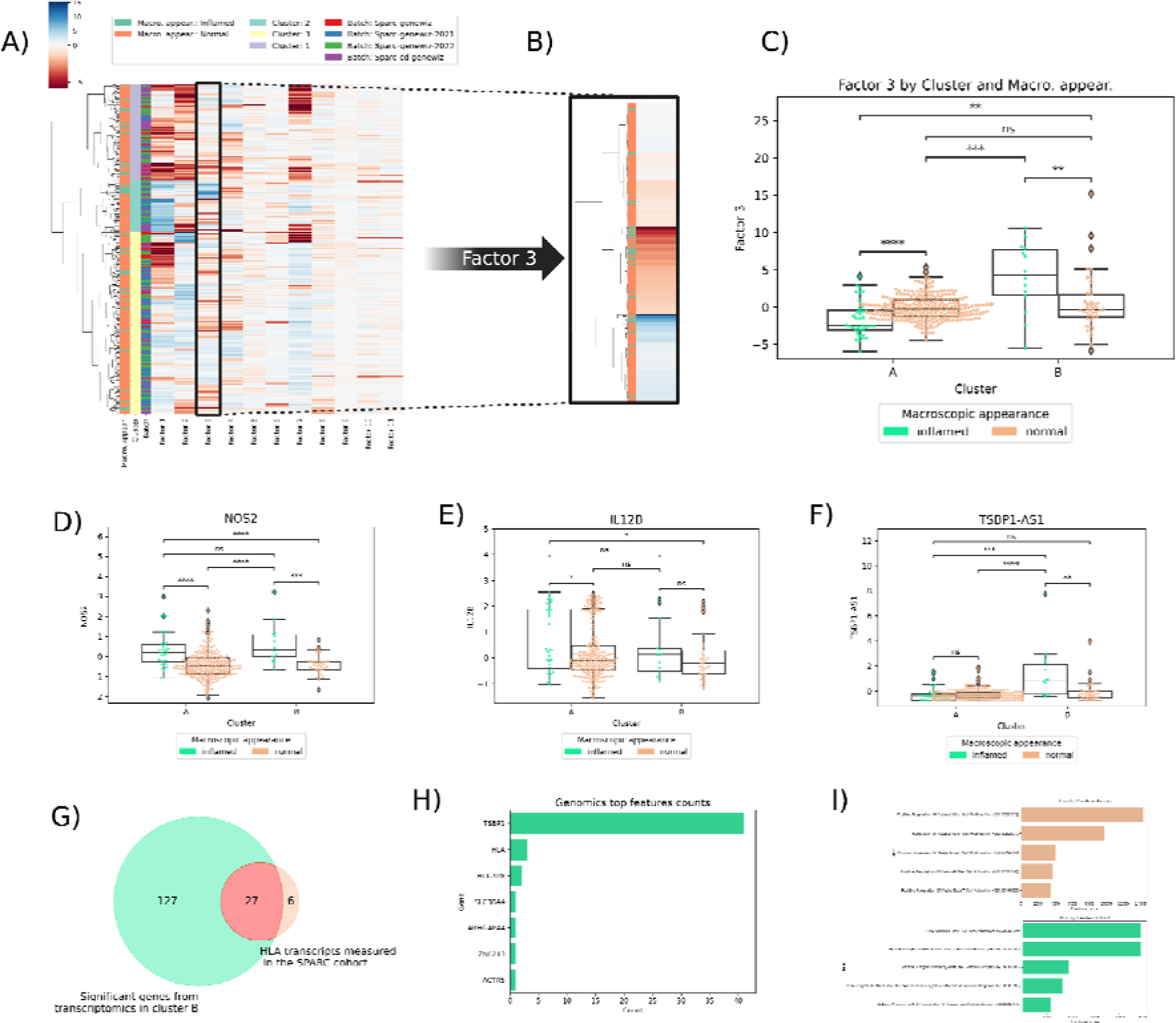
**A)** Hierarchical clustering applied to the factors from MOFA for multi-omics CD samples. Transcriptomics and proteomics data are derived from colon and plasma, respectively. Clustering on all factors reveals three main clusters. **B)** Hierarchical clustering applied exclusively to factor 3 and the reported macroscopic appearance for each sample (inflamed/normal). Samples with extreme values of factor 3 (dark-red and dark-blue) are likely to be inflamed, while values of factor 3 closer to zero (light-red, light-blue, and white) are less likely to be inflamed. **C)** Distribution of the factor 3 values for the two identified subpopulations (A: cluster 1 and 3, and B: cluster 2) stratified by macroscopic appearance (inflamed/normal). **D-F)** Distribution of the expression values for NOS2 (proteomics), IL12B (proteomics), and TSBP1-AS1 (transcriptomics) across the two subpopulations (A and B), stratified by macroscopic appearance. The three features are an example of an inflammation marker, cluster A specific marker, and cluster B specific marker, respectively. **G)** Proportion of HLA genes among the top 15 transcriptomic features in factor 3. **H)** Most common genes of the top 150 genomics features (SNPs). TSBP1 SNPs correspond to approximately 30% of the top genomics features. **I)** Top 5 enriched pathways using the significant proteomics features for the inflamed cluster A (orange) and significant transcriptomics features for the inflamed cluster B (green). Statistical significance is measured with Mann-Whitney-Wilcoxon test two-sided (*Legend*: ∗∗∗∗ = *p-*value < 0.0001; ∗∗∗ = *p-*value < 0.001; ∗∗ = *p-*value < 0.01; ∗∗ = *p-*value < 0.05; ns = non-significant).

After revealing the association between factor 3 and macroscopic appearance, we explored the distribution of factor 3 values across the three main identified clusters derived from the MOFA factors **(Figure 5B)**. Stratifying by the three clusters again suggests two subpopulations of inflamed samples, one comprising inflamed samples in cluster 1 and 3 (both present high negative factor 3 values), and one containing the inflamed samples of cluster 2 (positive factor 3 values). Therefore, we subsequently combined the inflamed samples of cluster 1 and 3 as they manifest a similar inflammation pattern based on factor 3. From this point on, the aggregate of clusters 1 and 3 is referred to as cluster A and cluster 2 as cluster B **(Figure 5C)**.

Next, we explored the top 150 features in each omics modality based on the absolute weights of factor 3. Our goal here was two-fold, i) identify which of these features can distinguish between inflamed and not inflamed samples and thus can be used as inflammation markers, and ii) investigate the different molecular signatures between the two potential subpopulations identified for inflamed samples. Among the top features in factor 3, we found several markers, both up- and down-regulated, in proteomics/transcriptomics that differentiate between inflamed and normal such as CXCL9, HLA-DRA, INHBB, SERPINA3, and NOS2 **(Figure 5D and Supplementary Figure 9)**. All of these proteomics or transcriptomics features have potential use as biomarkers for intestinal inflammation in the context of CD.

Similarly, to identify the features that characterize each of the two subpopulations of inflamed samples, we applied ANOVA to the top 150 proteomics and transcriptomics features and chi-square for the top genomics ones (*p*-value < 0.01), using cluster-A-inflamed vs rest and cluster-B-inflamed vs rest as groups. This revealed a subset of features characterizing the two subpopulations **(Figure 5E and F).** When investigating the transcriptomics features characteristic of cluster B, we found a large majority of these transcripts were Human Leukocyte Antigen (HLA) genes **(Figure 5G)**, a set of genes responsible for regulation of the immune system and involved in antigen processing and presentation **(Supplementary Figure 10)**. Furthermore, we found that one of the transcripts characteristic to cluster B, TSBP1-AS1 **(Figure 5F)**, corresponds to the most occurring SNPs highlighted in genomics, as 41 SNPs of this gene appear in the top 150 genomics features **(Figure 5H).** As a reference, there are 361 SNPs for this gene in the total 166,916 SNPs analyzed. In other words, while the SNPs of this gene represent 0.2% of the genomics features, it is over-represented among the top 150 genomics features in factor 3 with ∼27%. The SNPs of this antisense non-coding RNA (ncRNA) are also located in the HLA region in chromosome 6, suggesting an association between these SNPs and regulation of HLA genes. Cluster A, on the other hand, presented a smaller number of characteristic features. One of them is the IL12B in proteomics **(Figure 5E)**, a cytokine that acts on T and natural killer cells, and is up-regulated in cluster A.

Lastly, we investigated the pathways enriched using the characteristic features of the two inflamed subpopulations. Among the enriched pathways for proteomics, we found several inflammation-related pathways related to the innate immune system for cluster A (e.g., natural killer proliferation and activation) **(Figure 5I)**. In the case of transcriptomics pathways for cluster B, we observed pathways related to antigen processing and presentation, immunoglobulin production and the MHC class. Summarizing, our results suggest that the differentiation between these two subpopulations of inflamed samples is due to cluster B developing a more robust or heightened adaptive immune response characterized by high expression of HLA genes and other inflammation-related pathways.

## 4. Conclusion

In this study, we conducted a multi-omics analysis on samples from a large IBD patient cohort aimed at identifying distinct patient subgroups. We first assessed whether a ML classifier could accurately distinguish between UC and CD samples, by exclusively training it on multi-omics data. The performance of the model indicates that multi-omics can be used to accurately predict samples between the two conditions. Furthermore, we leveraged the interpretability of the model used to investigate its most predictive omics features. As expected, a large proportion of them have been previously reported in the literature and associated with either of the indications, thus serving as a control on the method. However, others have not yet been linked with IBD and should be investigated further as they might play a role in the pathophysiology of the disease and could be used as biomarkers for diagnosing patients with indeterminate colitis.

In parallel, we delved deeper into each IBD subtype —CD and UC— by categorizing patients into subgroups using MOFA (Multi-Omics Factor Analysis). This exploration unveiled distinct subpopulations within each disease category, offering promising avenues for the application of precision medicine strategies. Interestingly, a few omics signatures appear to be mainly responsible for said separation. Among these omics, we found previously associated markers with IBD such as IL17A, SERPINA3 and CXCL9 as well as novel genes such as TSBP1-AS1 and ZNF268. To summarize, our findings lay the groundwork for further research into the molecular underpinnings of IBD and highlight the potential for tailored therapeutic interventions based on specific omics profiles.

Several limitations must be acknowledged in the present study. Firstly, the absence of comprehensive omics data for all samples constrained the effective sample size, which limited the breadth of our analysis. Secondly, the ML classifier employed demonstrates significant accuracy in distinguishing between patient groups based on omics profiles; however, the performance is lower for UC. Finally, despite this study representing one of the largest patient-level investigations into IBD to date, the sample size could still benefit from expansion to enhance the robustness and generalizability of our findings.

In the future, we envision several potential extensions of our work. First, we aim at investigating additional omics modalities beyond the three analyzed to increase the breadth of molecular coverage and ultimately uncover new biological insights. To achieve this, untargeted LC-MS- and NMR-based metabolomics should be performed to add an extra layer of biology. Additionally, we aim at using multiple tissue information concurrently when training a classifier (e.g., concatenating multiple transcriptomics samples in patients with this information available). Finally, the strategy to address the missing values needs to be revisited with a less conservative approach, such that we do not simply use only the samples with all omics types available. Overall, we intend to re-assess our findings as patients in the SPARC cohort continue to be phenotyped. Additionally, it is necessary to confirm these findings in independent cohorts to ensure their generalizability. The performance of the ML model would also likely benefit from the aforementioned steps.

To summarize, our work identified biomarkers that i) can distinguish between CD and UC and ii) can distinguish between distinct IBD subpopulations: UC patients with higher versus lower disease severity and CD patients with distinct types of inflammation. Our findings will have a significant impact on clinical practice as they enable the identification of relevant patient subpopulations for which personalized treatment regimes can be developed.

## Data Availability

SPARC IBD is available upon approved application to Crohn’s & Colitis Foundation IBD Plexus (https://www.crohnscolitisfoundation.org/ibd-plexus). We released the scripts and notebooks to reproduce the work at https://github.com/enveda/sparc-multiomics.

## Acknowledgments

We would like to acknowledge Dr. David Healey and Dr. Biswapriya B. Misra for thoughtful comments and feedback on the manuscript.

The results published here are in whole from the Study of a Prospective Adult Research Cohort with IBD (SPARC IBD). SPARC IBD is a component of the Crohn’s & Colitis Foundation’s IBD Plexus data exchange platform. SPARC IBD enrolls patients with an established or new diagnosis of IBD from sites throughout the United States and links data collected from the electronic health record and study specific case report forms. Patients also provide blood, stool and biopsy samples at selected times during follow-up. The design and implementation of the SPARC IBD cohort has been previously described.

## Authors’ contributions

AJP, DDF, and KAW designed the study. AJP, SC, and DE prepared the datasets. AJP analyzed the datasets. APJ and DDF made the figures. AJP, MH, DDF, and KAW interpreted the results. AJP and DDF wrote the paper with help from KAW. All authors reviewed the manuscript. All authors have read and approved the final manuscript.

## Competing interests

All authors were employees of Enveda Biosciences Inc. during the course of this work and have real or potential ownership interest in the company.

## Notes

### Funding Statement

This study did not receive any funding

### Author Declarations

Crohn’s & Colitis Foundation (CCF)

