## Supplementary File for "Multi-omics data integration identifies novel biomarkers and patient subgroups in inflammatory bowel disease"

#### Supplementary Text 1: Details on data preparation

In the case of proteomics, this modality was generated using Olink® (Proximity Extension Assay (PEA)) on serum samples. For genomics, blood samples from patients in the SPARC cohort were genotyped with two technologies (Sazonovs *et al*., 2022): the Illumina Infinium Global Screening Array (GSAMD-24v1-0_20011747_A1) beadchip and whole-exome capture was performed with the Twist Custom Capture - 37Mb target (Twist Bioscience). Libraries were sequenced on the Illumina HiSeq platform according to standard protocols at Broad Institute's Genomics Platform (Cambridge, MA, USA). Lastly, for transcriptomics data, samples were collected as described in Raffals *et al.* (2022). rRNA depletion was performed with the QIAGEN FastSelect rRNA HMR Kit (Qiagen, Hilden, Germany), and strand-specific libraries were made with the NEBNext Ultra II Directional RNA Library Prep Kit (NEB, Ipswich, MA, USA). RNAseq was performed at GeneWiz with Hiseq 2x150bp on a HiSeq 4000 sequencing device. Sequence reads were trimmed to remove possible adapter sequences and nucleotides with poor quality using Trimmomatic v.0.36. The trimmed reads were mapped to the Homo sapiens GRCh38 reference genome available on ENSEMBL using the STAR aligner v.2.5.2b. Unique gene hit counts were calculated by using *featureCounts* from the Subread package v.1.5.2., and gene counts were calculated using *featureCounts* from the Subread package v1.5.2.

#### Supplementary Text 2: Merging different omics samples

The preparation of the data was heavily driven by the metadata and our downstream intent to deploy multi-omics approaches. This guided several steps in the processing, particularly when considering sample-wise processing **(see Figure below)**.

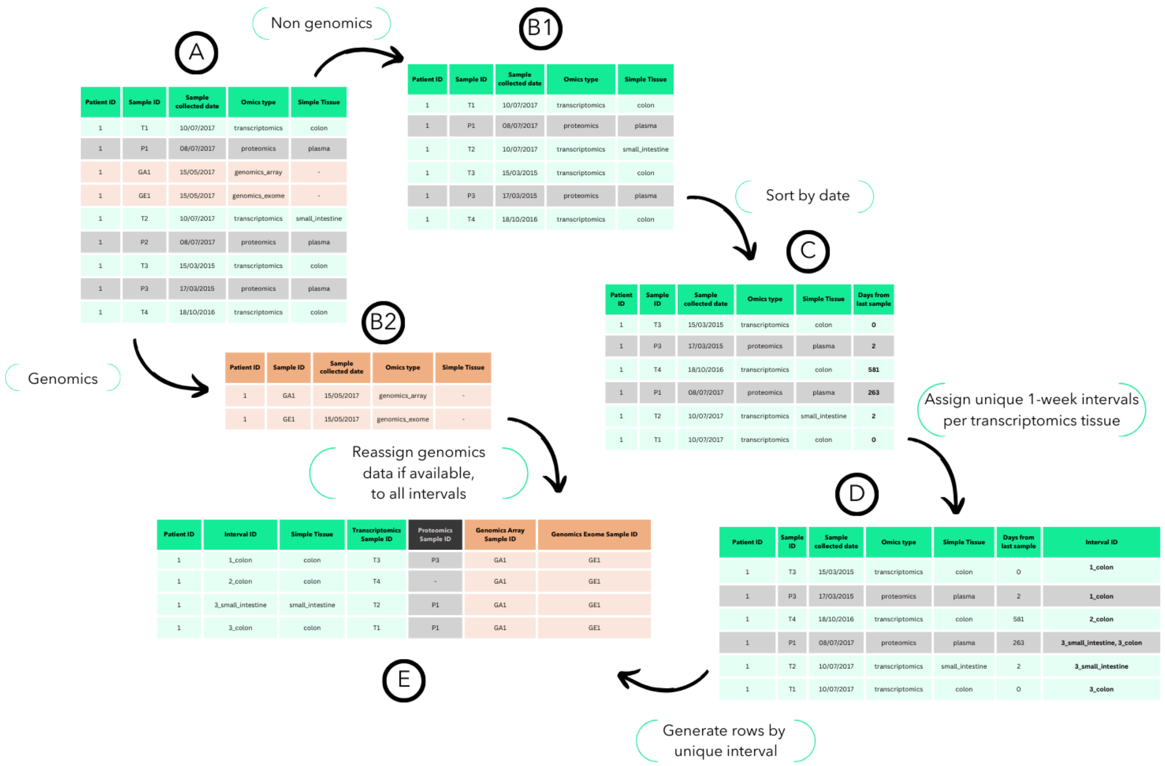


**Figure. Combining omics modalities across time intervals for a given patient.** Outline of the strategy to process the metadata with the goal of combining the different samples from a patient if they were taken within the same week and tissue. From the original long-format table (A), separating into non-genomics (i.e., transcriptomics and proteomics) (B1) and genomics information (B2); followed by sample collection date sorting (C), unique interval assignment (D), into final table generation (E) from collapsing omics sample IDs into one-week intervals.

To start off, the original data provided to us listed the metadata in the format of a table, where each sample is characterized by: (i) a patient identifier, which is unique per patient but not per sample; (ii) the sample identifier, which is unique for each sample and describes only one omics type at once, meaning each patient identifier can be mapped to multiple sample identifiers; (iii) the sample collected date, which is a timestamp for the time of collection of the sample; (iv) tissue information, which reports the tissue to which the sample belongs. (this is particularly relevant for transcriptomics, where we can have multiple omics types); (v) several other relevant variables such as macroscopic appearance, which are necessary for later steps but are not modified in the sample-wise metadata processing steps **(A)**.

In order to utilize the data, we need to match the multiple omics types such that each sample is representative of all available omics types in a time interval. For the length of the interval, we considered one week **(Supplementary Figure 1)**, as most of the samples for each interval were collected in this timespan and it is short enough to observe minimal phenotypic changes in most cases. Accordingly, we reorganized the metadata such that it exhibits the following characteristics: (i) an interval identifier, which reports for each patient, the interval of time considered (the same patient can have more than one valid interval of time); (ii) the interval identifier is also characterized by the tissue, since in the transcriptomics data, samples were at times retrieved from different tissues in a similar timespan; (iii) for each one week interval, we assigned the corresponding proteomics data, when available. All proteomics data pertains to the serum, meaning we can treat it as tissue independent; (iv) when available, for each unique patient, we assigned the genomics data, as we considered the time of collection for this data to be irrelevant; (v) for the variables that were not directly involved (such as macroscopic appearance), when in conflict across the multiple omics in the same interval, we privileged transcriptomics, as most of our variables of interest were stored in this data type to begin with.

### Supplementary Text 3: Multi-Omics Factor Analysis (MOFA)

The process behind MOFA involves the successive pruning of the input information until it is able to abstract biological coherence from the factors. This is driven by assigning the samples different biological profiles such that they are bundled in groups, which the model will then attempt to characterize via the factors. The features that contribute the most to these factors are thus more likely to be representative descriptors of the biological profile that the factor characterizes. This contribution can be assessed through the weights of each feature towards each factor. In the context of this study, the final input table was assigned each of the omics types (termed views, in accordance with the method), and considered the patient interval identifier as the samples and the macroscopic appearance as the sample groups. The views were assigned gaussian likelihoods. The explained variance used to drop features was 2%, as the authors describe in the original MOFA publication (Argelaguet *et al.* 2018). The model was then run with mostly default parameters, with an initial 25 factors that get pruned according to the explained variance. The sex (male/female) was used as a covariate. The trained models were saved after they converged. After reloading the factors from the respective models, we performed hierarchical clustering on the samples using euclidean distance, in order to categorize the patient intervals according to their omics profiles.

### Supplementary Figures

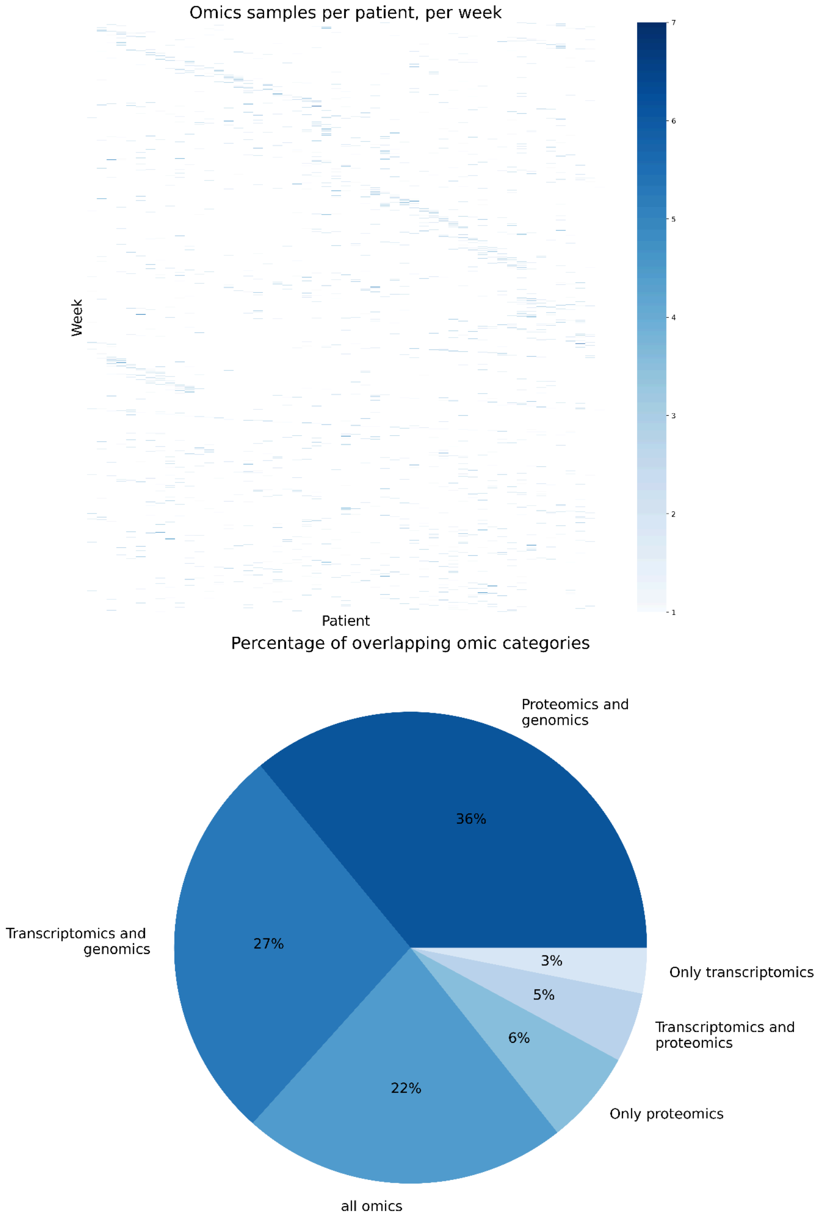


**Supplementary Figure 1**. Amount of omic samples per week (top). Percentage of overlapping omic types after the metadata rearrangement (bottom).

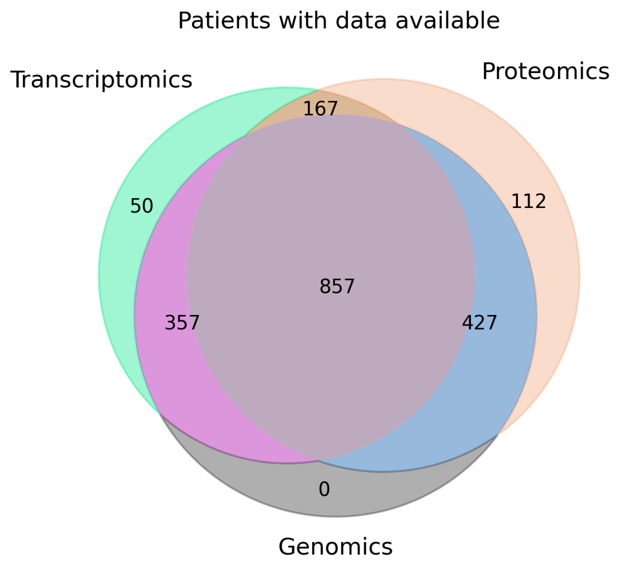


**Supplementary Figure 2**. Venn diagrams of patients overlap between all omics modalities.

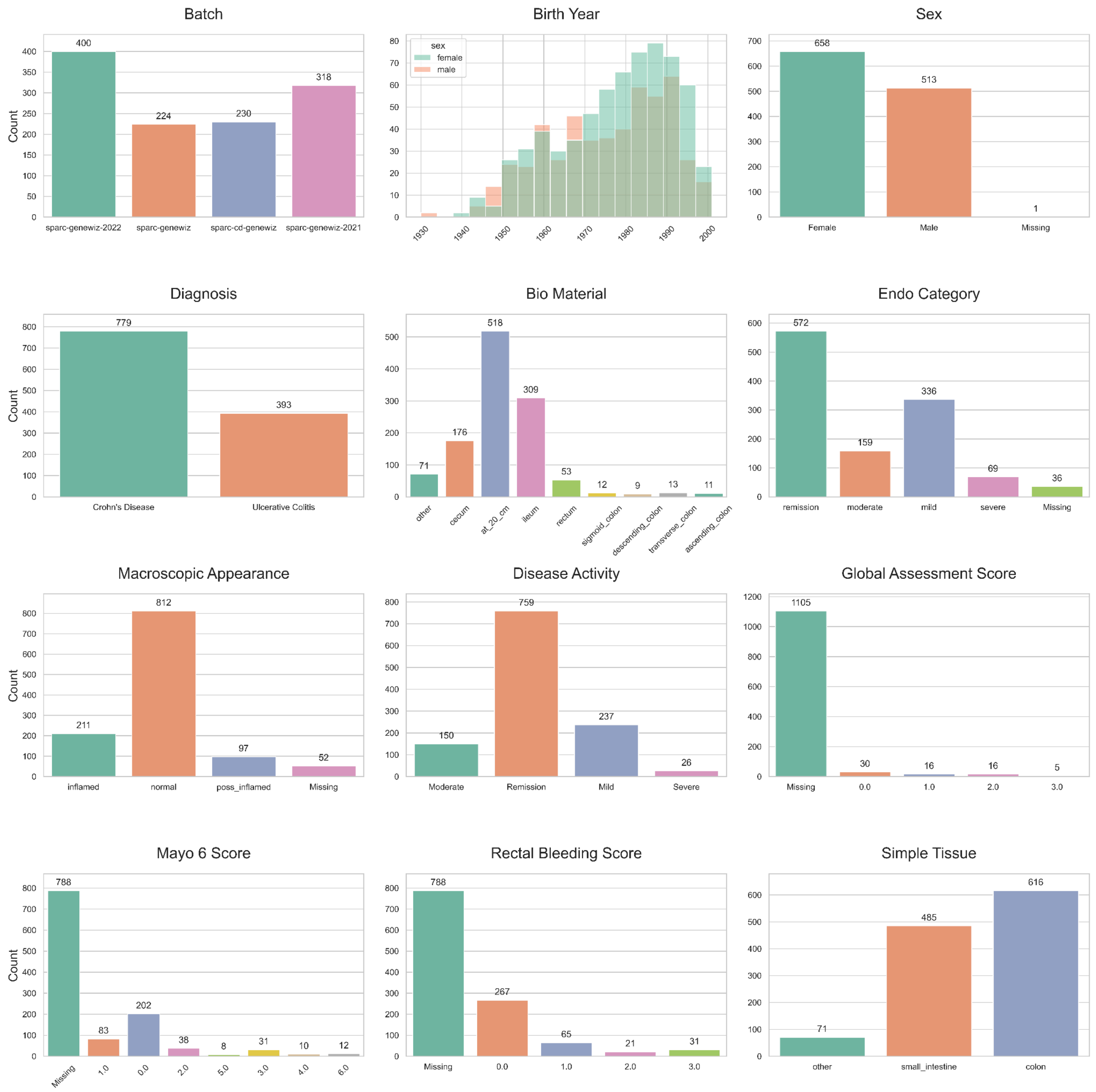


**Supplementary Figure 3.** Distribution of relevant metadata information in the SPARC dataset, subset to samples for which the patient has been diagnosed with UC or CD. These 1,023 samples from the 857 unique patients correspond to the ones used to train the ML classifier.

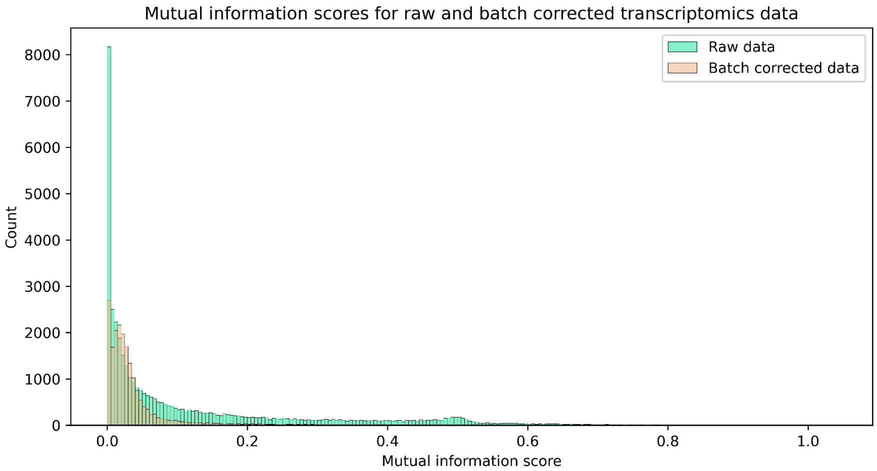


**Supplementary Figure 4.** Distribution of the MI scores for all batches for each gene in the raw transcriptomics data and after applying batch correction. The MI score measures the mutual dependence between the two variables, the higher the value, the stronger the relationship, with 0 representing no relationship. The plot shows how the distribution shifts to the left (less batch effect) when batch correction is applied.

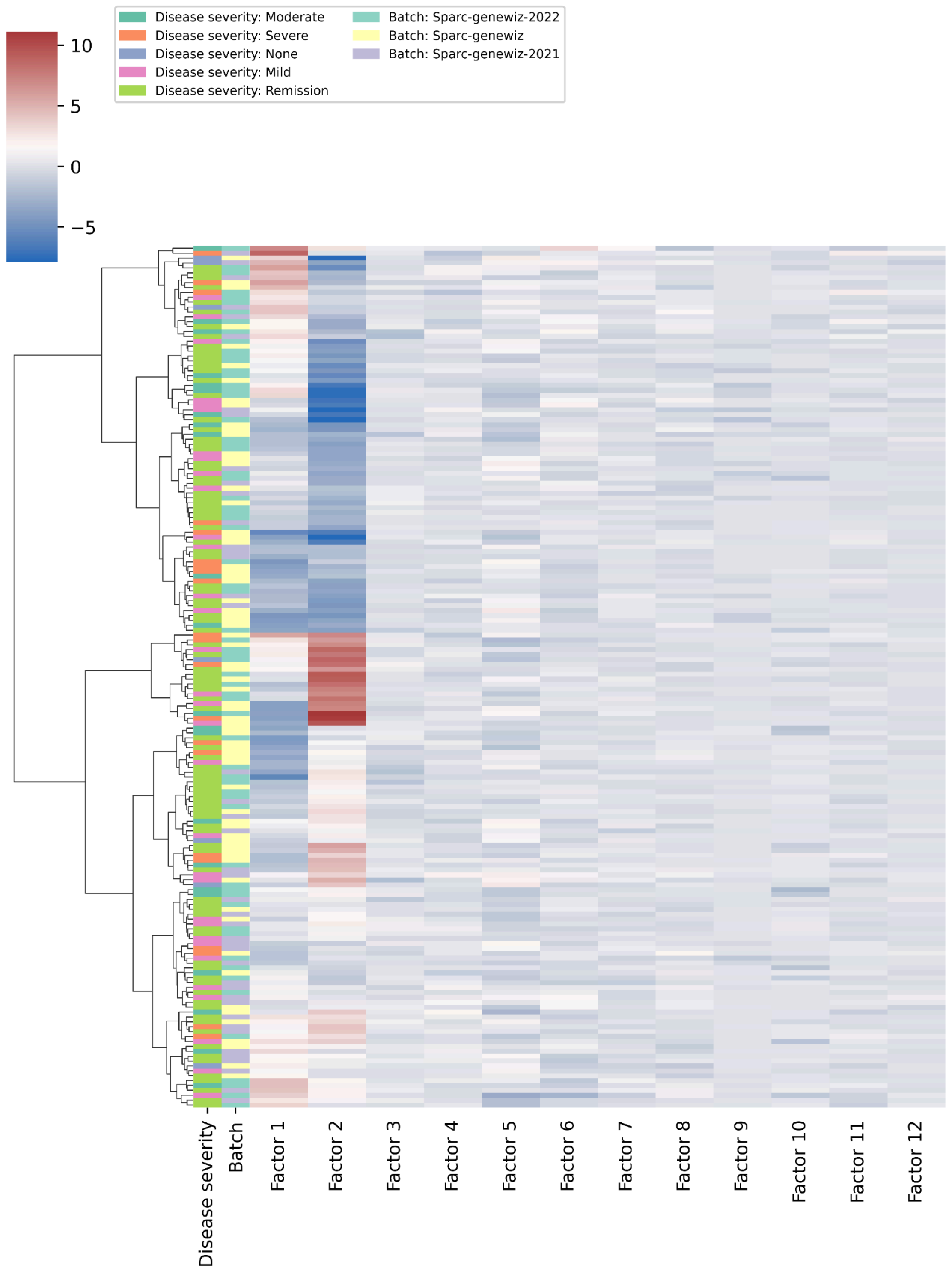


**Supplementary Figure 5.** Hierarchical clustering applied to the factors from MOFA for multi-omics UC samples. The first two columns indicate the disease severity and batch categories for each sample.

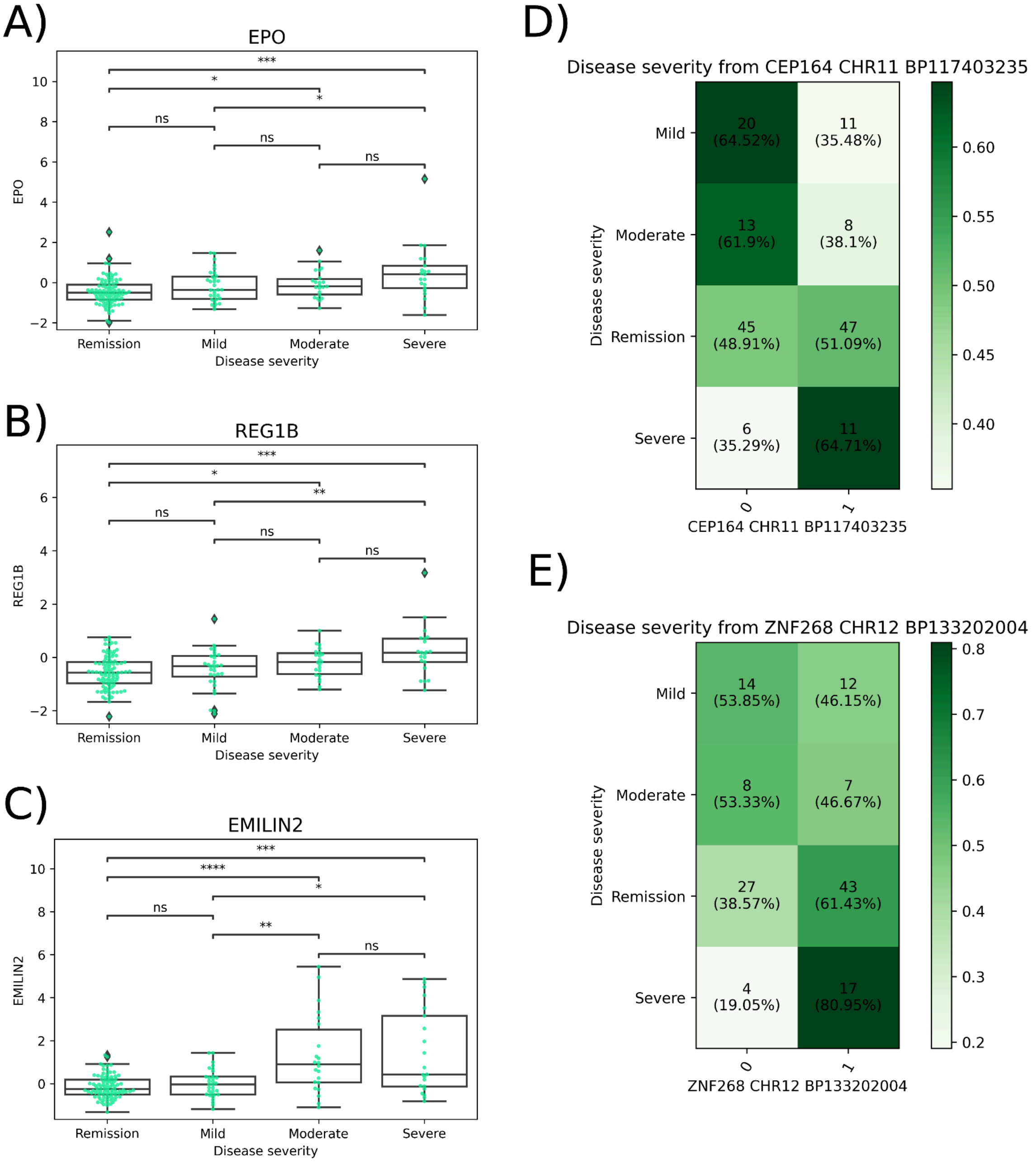


**Supplementary Figure 6.** Examples of features identified as significantly different between severe and mild and remission. (A) EPO (proteomics), (B), REG1B (proteomics) (C), EMILIN2 (transcriptomics), (D) Distribution of the genotypes (counts and percentage for each row) for ZNF268 (chr12_bp133202004) stratified by disease severity based on endoscopy. (E) Distribution of the genotypes (counts and percentage for each row) for CEP164 (chr11_bp117403235) stratified by disease severity based on endoscopy.

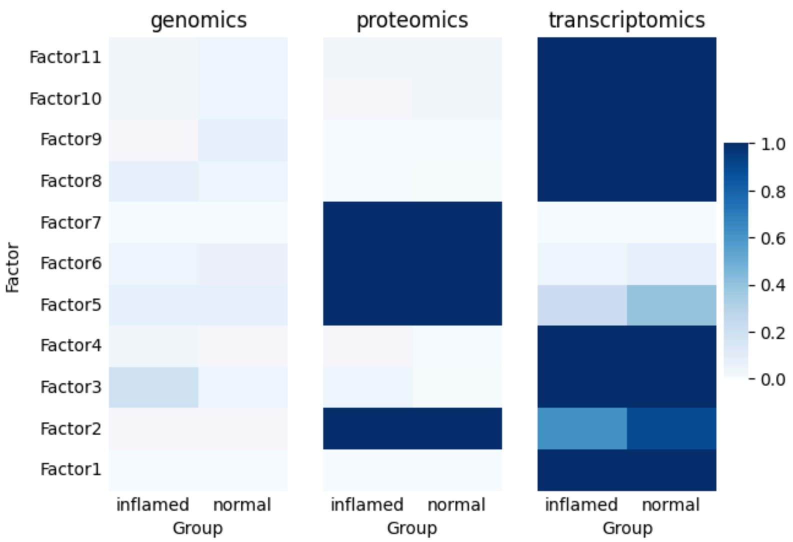


**Supplementary Figure 7.** R2 of the MOFA factors for CD colon by macroscopic appearance.

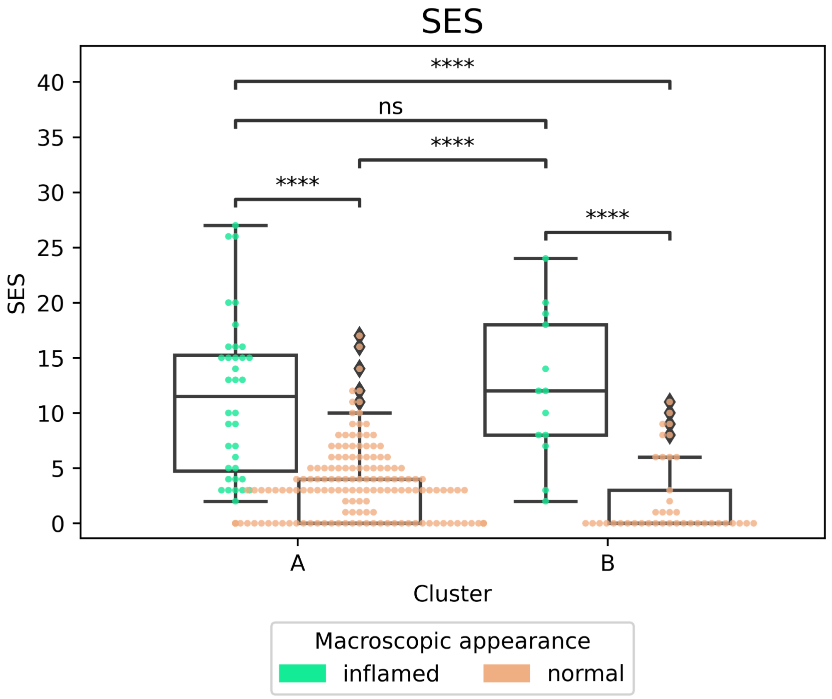


**Supplementary Figure 8.** Simple Endoscopic Score for Crohn's Disease (SES-CD) for each of the two clusters (A and B) stratifying by inflamed and normal samples.

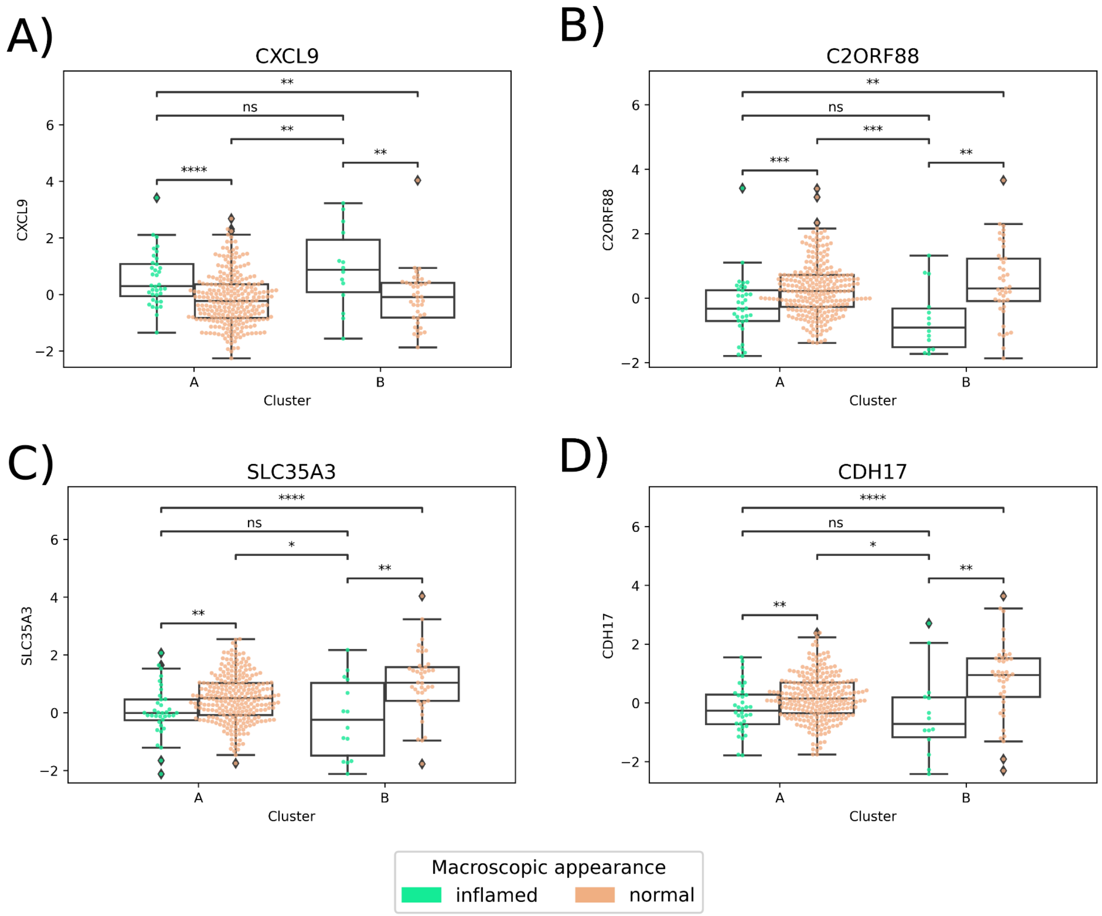


**Supplementary Figure 9.** Examples of genes identified significantly down-/up-regulated in inflamed samples. CXCL9 (proteomics) (A), C2orf88 (transcriptomics) (B), SLC35A3 (transcriptomics) (C) and SERPINA (transcriptomics) (D).

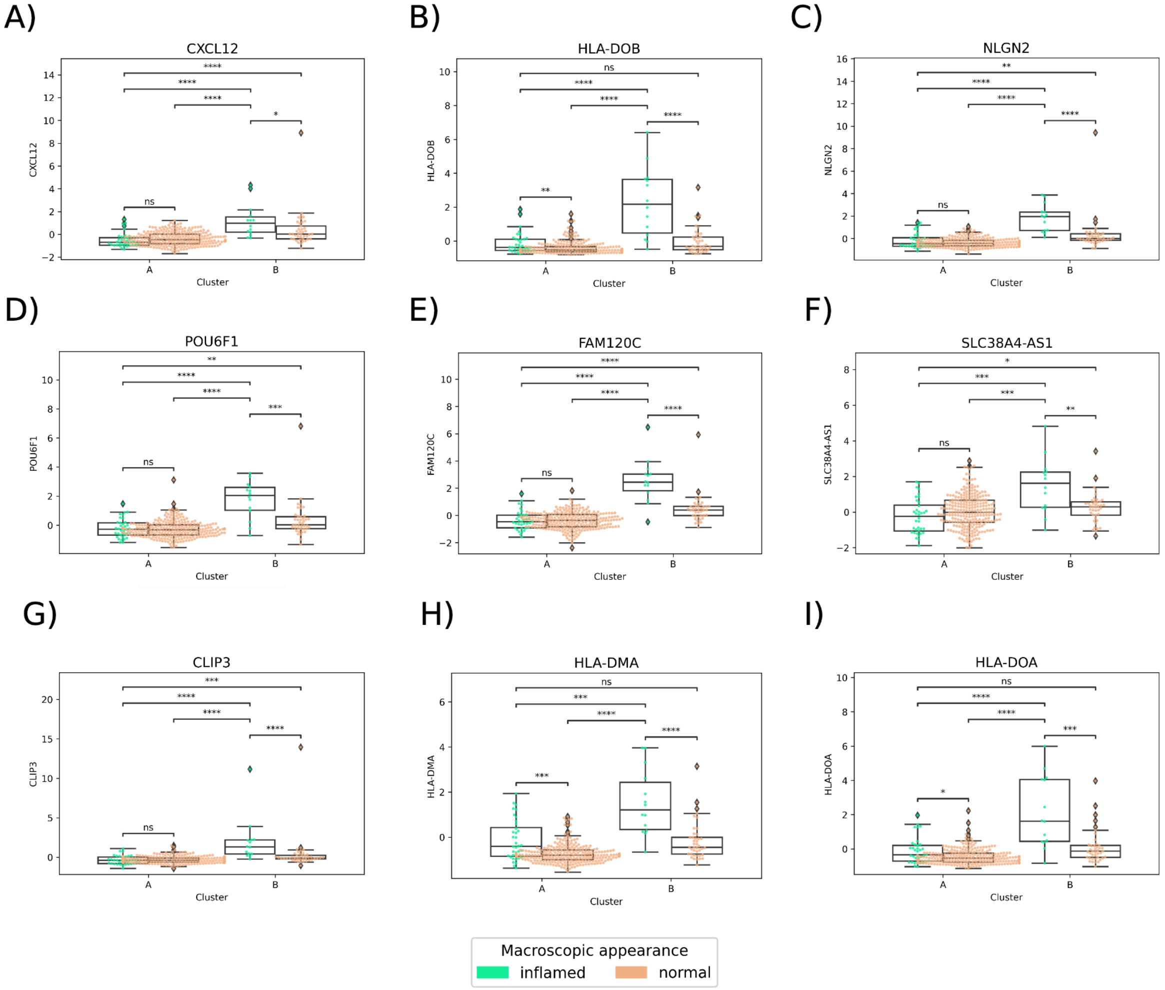


**Supplementary Figure 10.** Examples of genes identified significantly up-regulated in inflamed samples in Cluster B with respect to the rest, and thus, could be used as a biomarker for this subpopulation. Other features include ARL10, BOC, CH25H, CLIP3, CLSTN3, COL14A1, CX3CL1, CXCL12, DAZL, DLG4,DTX3, DZIP1, EBF1, EFS, ELN, EVC2, FAM120C, FGFR1, FLI1, FLRT2, GLI2, GLI3, GRID1, GRIK5, HLA-B, HLA-C, HLA-DMA, HLA-DMB, HLA-DOA, HLA-DOB, HLA-L, INPP4A, IRAG1, KCND1, KIAA1614, LSAMP, MAP3K12, MIR100HG, MOXD1, MPDZ, MYO5A, NCKAP5L, NLGN2, NR2F2-AS1, NXPH3, PACS1, PALD1, PHC1, POU6F1, PRDM6, PRELP, PTGDS, PTPRS, RAB3IL1, RBPMS, RECK, RGMA, RSPO1, RSPO3, RUNX1T1, SDC3, SLC22A17, SLC24A3, SLC27A1, SLC38A4-AS1, SOBP, SSC5D, ST8SIA1, STARD9, SYNGAP1, TCF7L1, TGFB1I1, TMCC2, TMEM200B, TNFSF12, TNS2, TRERF1, TSBP1-AS1, TTYH2, ZCCHC24, ZEB2, ZNF154, ZNF532, and PKIB.

### Supplementary Tables

| Parameter | Values |
|---|---|
| n_estimators | [**100**, 250, 500] |
| max_depth | [**3**, 5, 7] |
| eval_metric | [**logloss**, error, auc, aucpr] |

**Supplementary Table 1. Hyperparameter grid search used to train the XGBoost model.** The optimal parameters are highlighted in bold. The remaining parameters used are the default ones in the Python's *xgboost* library (v2.0.3).

| Variable | Category | Percentage of patients (n=537) |
|---|---|---|
| Omics available | Genomics | 100.00% (n=537) |
| | Proteomics | 100.00% (n=537) |
| | Transcriptomics | 100.00% (n=537) |
| | All (intersection) | 100.00% (n=537) |
| Sex | Male | 43.95% (n=236) |
| | Female | 55.87% (n=300) |
| | Undefined | 0.00% (n=1) |
| Diagnosis | UC | 35.94% (n=193) |
| | CD | 64.06% (n=344) |
| | Undefined | 0.00% (n=0) |
| Age | <18 | 0.00% (n=1) |
| | 18-25 | 10.62% (n=57) |
| | 25-35 | 28.12% (n=151) |
| | 35-45 | 23.09% (n=124) |
| | 34-55 | 16.95% (n=91) |
| | >55 | 13.78% (n=74) |

**Supplementary Table 2. Patient demographics of the SPARC IBD cohort used in our work.** We would like to note that the demographics reported here correspond to the patients for which the three omics modalities are available (see Supplementary Figure 1).
